# Clinical Features of Long COVID Patients Coinfected with *Mycoplasma pneumoniae*

**DOI:** 10.1101/2024.11.16.24317441

**Authors:** Xiaodan Zhu, Yanhua Li, Jinghua Wang, Weifei Gao

**Author notes:** Corresponding author Xiaodan Zhu.

## Abstract

**Background:** Since the SARS-CoV-2 pandemic, many patients have suffered prolonged complications, called “long COVID”. *Mycoplasma pneumoniae* is a common respiratory pathogen, Reports of simultaneous long COVID and *M. pneumoniae* infections are rare in the literature.

**Methods:** We analyzed the clinical data of patients with long COVID-19 who visited the Respiratory Clinic of The Affiliated Hospital of Hangzhou Normal University between January 1 and January 31, 2023, together with their laboratory and radiographic findings, with Pearson’s χ^2^ test.

**Results:** Fifty-two patients diagnosed with both long COVID and *M. pneumoniae* infection and 77 with long COVID only were compared. The ages, clinical symptoms, and comorbidities of the two groups did not differ significantly (P > 0.05). However, sex and imaging findings differed between the groups.

**Conclusions:** Our study showed that long COVID–*M. pneumoniae* coinfection was more commonly seen in females and patients with typical chest computed tomography images.

## 1. Introduction

By February 9, 2023, more than 755 million SARS-CoV-2 (COVID-19) infections and over 6.8 million COVID-associated deaths worldwide had been reported by the World Health Organization (WHO). Evidence quickly emerged that patients infected with COVID-19 not only experienced high levels of morbidity and mortality, but many (almost 70% of COVID-19 survivors) suffered a long period of complications. In October 2021, the term “long COVID” was introduced by WHO [1, 2], and applied to the persistent symptoms of COVID-19 survivors. According to WHO, symptom onset occurs during or up to more than 12 weeks after SARS-CoV-2 infection and can be explained by no alternative diagnosis. However, some experts consider that patients with long-term symptoms lasting 2 weeks have a mild disease and those with symptoms lasting 4 weeks have moderate-to-severe illness, whereas critically ill patients with symptoms lasting 6 weeks or more have “long COVID” syndrome [3]. Fatigue, cough, chest pain, dyspnea, palpitations, arthralgia, muscle pain, cognitive impairment, and anxiety are the main manifestations in patients with long COVID, and impose a substantial burden on the COVID survivor’s quality of life.

COVID-19 outbreaks often occur at the time of year when other respiratory infections are most common, and coinfections with other respiratory pathogens are a common feature of pandemics. *Mycoplasma pneumoniae* is a common atypical bacterium among respiratory pathogens [4, 5], and infections can occur in patients of all ages. It generally causes a mild self-limiting disease [6, 7]. However, when long COVID is complicated by coinfection with *M. pneumonia*, the treatment of COVID-19 becomes more difficult. The characteristics of patients with both long COVID and *M. pneumoniae* infection must be clarified to allow their clinical management. Long COVID complicated with *M. pneumoniae* infection is rarely documented in the medical community. A meaningful description of the clinical characteristics of patients with long COVID and *M. pneumoniae* coinfection, including laboratory results and radiological findings, is the aim of this study [8].

## 2. Methods

In this study, we retrospectively collected data on the presentations of long COVID patients, their demographic details, and test results from electronic records (laboratory data and chest computed tomography [CT] findings). Patients underwent the following laboratory tests: complete blood counts, C-reactive protein, creatine kinase, lactate dehydrogenase, and transaminase. The patients were diagnosed with long COVID and/or *M. pneumoniae* between January 1 and January 31, 2023, at the Respiratory Clinic at The Affiliated Hospital of Hangzhou Normal University(Hangzhou City, Zhejiang Province, China).

After the initial detection of COVID-19 antigen, the viral load becomes undetectable[16]. This marks the start of long COVID. Presentation with acute respiratory symptoms, such as fever and cough, and no viral antigen after its initial detection were used as the diagnostic criteria for long COVID. The diagnostic criterion for detecting Mycoplasma pneumoniae (M. pneumoniae) infections involves utilizing a method from a Japanese Antibody Test Kit designed to identify MP-IgM antibodies specific to M. pneumoniae.Colloidal gold immunochromatographic assay for rapid detection of specific anti-Mycoplasma pneumoniae IgM antibodies.These antibodies can typically be detected 5-6 days after infection. A titer of 1:160 or higher is considered positive. A positive result suggests a possible Mycoplasma pneumoniae infection, while a negative result does not completely rule out MP infection. And this method is particularly suitable for rapid screening. For confirm *M. pneumoniae* infection.It’s essential to conduct a comprehensive analysis in conjunction with clinical and imaging characteristics. [9, 10, 25].

## 3. Statistical analysis

We used SPSS ver. 25 (IBM SPSS) to analyze the data statistically. Categorical variables and continuous variables were summarized as frequencies or percentages and are expressed as means ± standard deviations (SD). Categorical variables were compared with Pearson’s χ^2^ test. A P value < 0.05 was considered significant [11].

## 4. Results

### 4.1 Clinical characteristics of participants

The data for 77 patients with long COVID and 52 diagnosed with both long COVID and *M. pneumoniae* infection were analyzed. All patients suffered mild acute COVID. The total number of long COVID patients included in the study was 129, 52 (40.31%) of whom men and 77 (59.69%) women. The mean age of the patients was 42.89 ± 15.89 years (range 16–88years). The two groups did not differ significantly in terms of age (P > 0.05), but differed significantly in the distribution of sex (P < 0.05). Female long-COVID patients were more frequently infected with *M. pneumonia* than male patients [odds ratio (OR) = 2.28]. Long COVID patients coinfected with *M. pneumoniae* were more likely to have symptoms such as cough, sputum production, palpitation, chest tightness, shortness of breath, or chest pain than were those not infected with *M. pneumonia* but these differences were not statistically significant (P > 0.05). Thirty-six (27.90%) patients enrolled in the study also suffered chronic conditions, such as asthma, hypertension, diabetes mellitus, anemia, and atrial fibrillation. However, there was no meaningful difference in comorbidities between the two groups (Table 1).

**Table 1.** The difference in clinical characteristics between long COVID patients and MP co-infected patients.

|  | Total | long COVID | long COVID Coinfected MP | P value |
| --- | --- | --- | --- | --- |
| **Patients (n)** | 129 | 77 | 52 |  |
| **Sex** |  |  |  | **0.029** |
| Male | 52 | 37 | 15 |  |
| Female | 77 | 40 | 37 |  |
| **Age (years)** |  |  |  | **0.056** |
| ≤20 | 2 | 1 | 1 |  |
| 21–40 | 67 | 40 | 27 |  |
| 41–60 | 37 | 17 | 20 |  |
| >61 | 23 | 19 | 4 |  |
| **Symptom** |  |  |  |  |
| Cough | 120 | 74 | 46 | 0.095 |
| Expectoration | 80 | 50 | 30 | 0.406 |
| palpitation | 10 | 6 | 4 | 0.983 |
| Chest pain | 7 | 3 | 4 | 0.351 |
| Chest tightness | 35 | 23 | 12 | 0.395 |
| Shortness of breath | 27 | 15 | 12 | 0.622 |
| **Chronic disease** | 36 | 23 | 13 | 0.205 |
| **CT scan result** |  |  |  | **0.032** |
| Ground-glass opacity (GGO) | 82 | 55 | 27 |  |
| No obvious abnormalities | 33 | 15 | 18 |  |

### 4.2 Comparisons of laboratory results of long COVID patients with or without *M. pneumonia* infection

A comparison of the laboratory test results of the long COVID patients and those coinfected with *M. pneumoniae* indicated that the white blood cell (WBC), red blood cell (RBC), hemoglobin, creatine kinase, and C-reactive protein levels did not differ significantly between the two groups. The patients with only long COVID had almost normal laboratory parameters. However, increases in aspartate transaminase (AST) and alanine transaminase (ALT) were observed more frequently in long COVID patients without *M. pneumoniae* infection (Table 2).

**Table 2.** The difference in laboratory tests between long COVID patients and MP co-infected patients.

|  | Total | long COVID | long COVID<br>Coinfected MP |
| --- | --- | --- | --- |
| WBC (10<sup>9</sup> /L) | 6.81±1.84 | 6.92±1.92 | 6.63±1.71 |
| RBC (10<sup>12</sup>/L) | 4.66±0.50 | 4.73±0.50 | 4.56±0.48 |
| PLT (10<sup>9</sup> /L) | 246±57 | 242±51 | 251±64 |
| Hb (g/L) | 135±17 | 139±16 | 134±16 |
| CRP >10 mg/L | 2.77±2.86 | 2.78±2.50 | 2.76±3.54 |
| LDH (U/L) | 195±41 | 198±45 | 191±31 |
| CK (U/L) | 86±38 | 92±41 | 75±29 |
| CK-MB(ng/ml) | 2.05±0.83 | 2.06±0.69 | 2.03±1.07 |
| ALT (U/L) | 29±21 | 32±24 | 22±14 |
| AST (U/L) | 26±11 | 29±13 | 21±3 |

### 4.3 Chest CT findings

Fourteen patients were not examined with CT. Fifty-five patients in the long COVID group (55/70, 78.57%) had positive CT findings, whereas 60% (27/45) of long COVID patients coinfected with *M. pneumoniae* had positive imaging findings. These findings did differ significantly (P = 0.032). Abnormal imaging was more commonly seen in the of long COVID patients than in the long COVID patients coinfected with *M. pneumoniae* (OR =2.44). Ground glass opacity (GGO) distributed in the dorsal outer zone was the predominant abnormality detected in these patients (Figure 1) [12].

**Figure 1.**
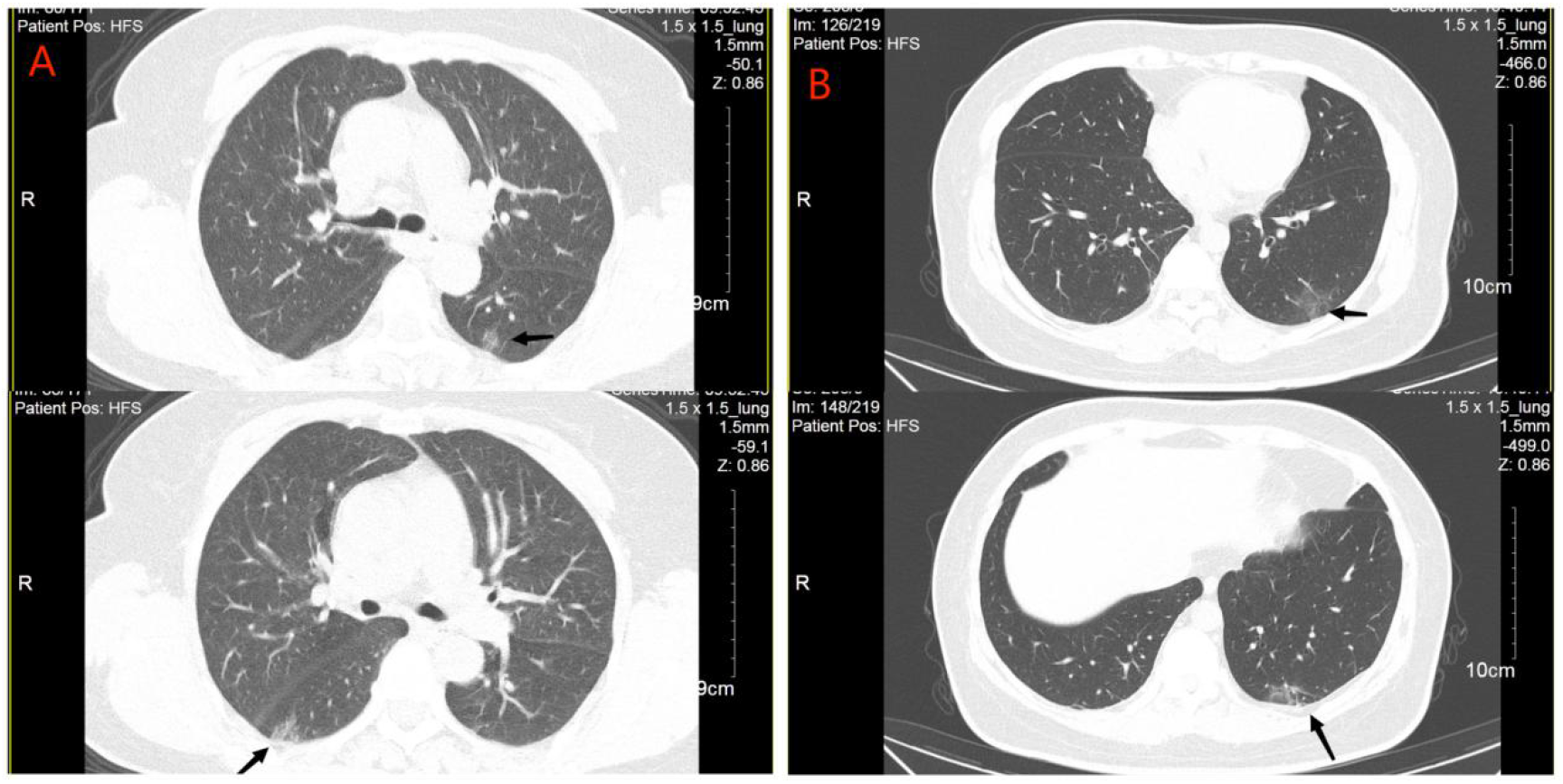
A: The CT examination of long COVID patients with M. pneumoniae infection demonstrated a GGO in the dorsal outer zone of the bilateral lung(arrowhead); B: The CT examination of long COVID patients with M. pneumoniae infection showed a GGO in the dorsal outer zone of the left lower lobe(arrowhead).

## 5. Discussion

Social distance keeping and mandatory mask-wearing have led to a significant decrease in the incidence of various respiratory infectious diseases, including Mycoplasma pneumoniae and influenza. Because of the lifting of COVID-19 restrictions, China’s epidemiological data on M pneumoniae is increasing.

Coinfections with coronaviruses and other respiratory pathogens have frequently been observed since the the lifting of COVID-19 restrictions. Several studies of these coinfections have been reported. Lansbury et al. [13] detected a bacterial coinfection in 7% of hospitalized COVID-19 patients, and *M. pneumonia* was the most common bacterium involved. Tang et al. [14] reported that coinfection correlated with elevated procalcitonin(PCT) levels in patients with COVID-19 pneumonia, which was successfully treated with anti-inflammatory therapies. However, there have been few comprehensive studies of long COVID patients with coinfections.

### 5.1 Risk factors of long COVID with *M. pneumoniae* coinfection

In this study, a high proportion (42.3%) of long COVID patients was coinfected with *M. pneumonia*, and coinfection was detected significantly more often in women than men. Several papers have suggested that underlying immune-system disorders cause the persistent symptoms of long COVID. High levels of autoantibodies have been found in some patients with COVID-19 more generally.Several studies have indicated that low or no production of SARS-CoV-2 antibodies and other insufficient immune responses during the acute phase of COVID-19 can predict the development of long COVID[26,27].The innate and adaptive immune responses could be the essential factors making females more susceptible to long COVID [15].Women being more likely to have lower levels of antibodies and being less likely to seroconvert overall [28].

The primary clinical symptoms of long COVID are fatigue, chest pain, dyspnea, palpitations, arthralgia, muscle pain, cognitive impairment, and anxiety. In recent studies, fatigue was the most widespread symptom in long COVID patients. And cognition-related symptoms and cardiovascular complications were also reported in the latest international survey[3].Because our data were all collected from respiratory clinic patients, respiratory dysfunction was recorded most frequently. Cough was the main symptom for which long COVID patients visited the clinic, followed by expectoration, shortness of breath, and chest tightness. These respiratory symptoms may be related to virus-induced changes in lung volume and airway clearance.Patients with *M. pneumoniae* also reported non-specific respiratory symptoms, including general malaise and dry cough. The presence of mycoplasma coinfection could be easily overlooked due to the similar presentations[29].

Koc et al. [16] reported that bronchial asthma, type 2 diabetes, and psychological disorders increase the risk of long COVID. Patients with hypertension, diabetes, and anxiety were also included in the present study. The risk of long COVID coinfected with *M. pneumoniae* was not increased by comorbidities.

### 5.2 Clinical indicators in long COVID patients coinfected with *M. pneumoniae*

Yong et al. [17] reported that patients with long COVID had higher levels of C-reactive protein, leukocytes, lymphocytes, and lactate dehydrogenase than those without long COVID. However, we detected the expected levels of leukocytes, erythrocytes, platelets, hemoglobin, C-reactive protein, lactate dehydrogenase, creatine kinase, and CK-MB in both long COVID patients with and without *M. pneumoniae* coinfection. The significant deviations from normal observed in inflammatory and vascular biomarker levels may be attributable to the severity of the COVID infection. The patients enrolled in our study were all mildly ill. A low viral load is likely to trigger a lower grade inflammatory response in the acute phase of viral infection, resulting in less damage to the host during long COVID.

### 5.3 Imaging findings of long COVID patients coinfected with *M. pneumoniae*

Computed tomography is the most widely used imaging technology for respiratory diseases [18, 19]. The chest CT manifestations in adults with COVID-19 or *M. pneumoniae* are the interstitial changes [20]. Ground-glass opacity (GGO), consolidation, linear opacity, vacuolar sign, and a crazy-paving pattern are common pulmonary lesions in COVID-19 patients. In contrast, the lesions in *M. pneumonia*-infected patients are predominantly distributed along the bronchi, and the bronchial wall is thickened, accompanied by tree buds/fog signs [21]. Our study showed that in patients with long COVID with or without *M. pneumoniae* infection, the major imaging change was GGO, distributed in the dorsal outer zone.

*Mycoplasma pneumoniae* infection is not considered to pose a risk of lung lesions in patients with long COVID. However, Bazdar et al. [22] reported 29 different imaging findings in long COVID patients, classified as interstitial (fibrotic), pleural, airway, and other parenchymal abnormalities. While CT imaging can provide valuable insights into the physiological effects of Long COVID, the condition’s presentation can be highly variable among patients. Imaging findings should be interpreted with clinical symptoms and other diagnostic information.To properly evaluate long COVID, more research is required into the lung imaging findings of these patients [23].

## 6. Conclusions

In this study, we detected no association between *M. pneumoniae* coinfection and biochemical abnormalities in patients with long COVID. Female long COVID patients and those with normal radiography were significantly more likely to present with *M. pneumoniae* coinfection. No correlation detected was between *M. pneumoniae* coinfection in long COVID patients and their pre-existing conditions, age, or symptoms. Various guidelines focus on the treatment and management of COVID-19, but there is little information on long COVID in these guidelines [24]. Based on the results of our study, the clinical symptoms of the syndrome were the primary health problem and additional antibiotics for *M. pneumoniae* coinfection may be not necessary. Improving the clinical characterization of long COVID may be critical to the provision of adequate treatment options.

## Data Availability

All data produced in the present study are available upon reasonable request to the authors

## 7. Acknowledgments

We would like to thank all our colleagues involved in this project for their timely assistance. This work was supported by the Affiliated Hospital of Hangzhou Normal University.

## 8. Authors’Contributions

Xiaodan conceptualized the study, Jinghua were involved in the design, and analysis of the study. Xiaodan drew up the manuscript and it was revised by the other authors. The final manuscript was read and approved by all authors.

## References

1. Davis B, Rothrock AN, Swetland S, et al. Viral and atypical respiratory co-infections in COVID-19: a systematic review and meta-analysis. J Am Coll Emerg Physicians Open. 2020 Jun 19;1(4):533–548. doi: 10.1002/emp2.12128.

2. Castanares-Zapatero D, Chalon P, Kohn L, et al. Pathophysiology and mechanism of long COVID: a comprehensive review. Ann Med. 2022 Dec;54(1):1473–1487. doi: 10.1080/07853890.2022.2076901.

3. Asadi-Pooya AA, Akbari A, Emami A,et al. Long COVID syndrome-associated brain fog. J Med Virol. 2022 Mar;94(3):979–984. doi: 10.1002/jmv.27404.

4. Fan BE, Lim KGE, Chong VCL, et al. COVID-19 and mycoplasma pneumoniae coinfection. Am J Hematol. 2020 Jun;95(6):723–724. doi: 10.1002/ajh.25785.

5. Ito K, Yokoyama T, Horiuchi M, et al. COVID-19 pneumonia suspected to be co-infection with Mycoplasma pneumoniae and improved by early administration of favipiravir and ciclesonide. Respirol Case Rep. 2021 Aug 3;9(9):e0821. doi: 10.1002/rcr2.821.

6. Chen FL, Wang CH, Hung CS, et al. Co-infection with an atypical pathogen of COVID-19 in a young. J Microbiol Immunol Infect. 2021 Feb;54(1):154–155. doi: 10.1016/j.jmii.2020.05.007.

7. Zha L, Shen J, Tefsen B, et al. Clinical features and outcomes of adult COVID-19 patients co-infected with Mycoplasma pneumoniae. J Infect. 2020 Sep;81(3):e12–e15. doi: 10.1016/j.jinf.2020.07.010.

8. Crook H, Raza S, Nowell J, et al. Long covid-mechanisms, risk factors, and management. BMJ. 2021 Jul 26;374:n1648. doi: 10.1136/bmj.n1648.

9. Miyashita N, Nakamori Y, Ogata M, et al. Clinical Differences between Community-Acquired Mycoplasma pneumoniae Pneumonia and COVID-19 Pneumonia. J Clin Med. 2022 Feb 12;11(4):964. doi: 10.3390/jcm11040964.

10. Cho YJ, Han MS, Kim WS, et al. Correlation between chest radiographic findings and clinical features in hospitalized children with Mycoplasma pneumoniae pneumonia. PLoS One. 2019 Aug 28;14(8):e0219463. doi: 10.1371/journal.pone.0219463.

11. Cunha BA. The atypical pneumonias: clinical diagnosis and importance. Clin Microbiol Infect. 2006 May;12 Suppl 3:12–24. doi: 10.1111/j.1469-0691.2006.01393.x.

12. Liu J, Wang Y, He G, et al. Quantitative CT comparison between COVID-19 and mycoplasma pneumonia suspected as COVID-19: a longitudinal study. BMC Med Imaging. 2022 Feb 6;22(1):21. doi: 10.1186/s12880-022-00750-4.

13. Lansbury L, Lim B, Baskaran V, et al. Co-infections in people with COVID-19: a systematic review and meta-analysis. J Infect. 2020 Aug;81(2):266–275. doi: 10.1016/j.jinf.2020.05.046.

14. Tang ML, Li YQ, Chen X, et al. Co-Infection with Common Respiratory Pathogens and SARS-CoV-2 in Patients with COVID-19 Pneumonia and Laboratory Biochemistry Findings: A Retrospective Cross-Sectional Study of 78 Patients from a Single Center in China. Med Sci Monit. 2021 Jan 3;27:e929783. doi: 10.12659/MSM.929783.

15. Sylvester SV, Rusu R, Chan B, et al. Sex differences in sequelae from COVID-19 infection and in long COVID syndrome: a review. Curr Med Res Opin. 2022 Aug;38(8):1391–1399. doi: 10.1080/03007995.2022.2081454.

16. Koc HC, Xiao J, Liu W, et al. Long COVID and its Management. Int J Biol Sci. 2022 Jul 11;18(12):4768–4780. doi: 10.7150/ijbs.75056.

17. Yong SJ, Halim A, Halim M, et al. Inflammatory and vascular biomarkers in post-COVID-19 syndrome: A systematic review and meta-analysis of over 20 biomarkers. Rev Med Virol. 2023 Mar;33(2):e2424. doi: 10.1002/rmv.2424.

18. Chaudhry R, Sreenath K, Vinayaraj EV, et al. Mycoplasma pneumoniae co-infection with SARS-CoV-2: A case report. Access Microbiol. 2021 Mar 10;3(3):000212. doi: 10.1099/acmi.0.000212.

19. Wang X, Wang Z, Sun Z, et al. Comparison of Clinical and Chest CT Characteristics of Adult Patients with COVID-19 and Mycoplasma Pneumonia. Iran J Radiol. 2021 Jan 13;18(1):e106612. 10.5812/iranjradiol.106612.

20. Alqahtani JS, Alghamdi SM, Aldhahir AM, et al. Thoracic imaging outcomes in COVID-19 survivors. World J Radiol. 2021 Jun 28;13(6):149–156. doi: 10.4329/wjr.v13.i6.149.

21. Gayam V, Konala VM, Naramala S, et al. Presenting characteristics, comorbidities, and outcomes of patients coinfected with COVID-19 and Mycoplasma pneumoniae in the USA. J Med Virol. 2020 Oct;92(10):2181–2187. doi: 10.1002/jmv.26026.

22. Bazdar S, Kwee AKAL, Houweling L, et al. A Systematic Review of Chest Imaging Findings in Long COVID Patients. J Pers Med. 2023 Feb 1;13(2):282. doi: 10.3390/jpm13020282.

23. Huo X, Xue X, Yuan S, et al. [Early differential diagnosis between COVID-19 and mycoplasma pneumonia with chest CT scan]. Zhejiang Da Xue Xue Bao Yi Xue Ban. 2020 Aug 25;49(4):468–473. Chinese. doi: 10.3785/j.issn.1008-9292.2020.07.04.

24. Sykes DL, Holdsworth L, Jawad N, et al. Post-COVID-19 Symptom Burden: What is Long-COVID and How Should We Manage It? Lung. 2021 Apr;199(2):113–119. doi: 10.1007/s00408-021-00423-z.

25. Guarino M, Perna B, Cuoghi F, et al. Role of Intracellular Pulmonary Pathogens during SARS-CoV-2 Infection in the First Pandemic Wave of COVID-19: Clinical and Prognostic Significance in a Case Series of 1200 Patients. Microorganisms. 2022 Aug 12;10(8):1636. doi: 10.3390/microorganisms10081636.

26. García-Abellán, J., Padilla, S., Fernández-González, M.,et al. Antibody Response to SARS-CoV-2 is Associated with Long-term Clinical Outcome in Patients with COVID-19: a Longitudinal Study. Journal of clinical immunology, 2021;41(7), 1490–1501.

27. Augustin, M., Schommers, P., Stecher, M., et al. Post-COVID syndrome in non-hospitalised patients with COVID-19: a longitudinal prospective cohort study. The Lancet regional health. Europe, 2021;6, 100122.

28. Davis, H. E., McCorkell, L., Vogel, J. M., & Topol, E. J. Long COVID: major findings, mechanisms and recommendations. Nature reviews. Microbiology, 2023;21(3), 133–146.

29. Huang, A. C., Huang, C. G., Yang, C. T., & Hu, H. C. Concomitant infection with COVID-19 and Mycoplasma pneumoniae. Biomedical journal, 2020;43(5), 458–461.

